# The Midnight Gap: Nighttime is associated with detrimental outcome in out-of-hospital cardiac arrests in Poland

**DOI:** 10.1101/2025.05.09.25327222

**Authors:** Anna Żądło, Monika Bednarek-Chałuda, Izabela A. Karpińska, Grzegorz Cebula, Tomasz Tokarek

## Abstract

**Objective:** Out-of-hospital cardiac arrest (OHCA) has low survival rates with worse outcomes at night due to delayed emergency medical services (EMS) response, resource limitations, and workforce fatigue. Timely resuscitation is crucial, but logistical challenges exacerbate disparities. Since randomized trials are unfeasible, all-comers registries provide essential data to bridge evidence gaps and improve EMS protocols. This study aimed to investigate the impact of day versus night shifts on OHCA outcomes, focusing on ROSC rates, 30-day survival, and timing metrics within EMS operations.

**Methods:** This study analyzed OHCA cases in Poland from September to November 2022 using paramedics records and national death registry data. Patients were grouped by time of cardiac arrest (on-hours: 7:00 AM–6:59 PM; off-hours: 7:00 PM–6:59 AM) and matched 1:1 using propensity score analysis (1194 pairs).

**Results:** Our findings revealed significant disparities in OHCA outcomes between day and night shifts. ROSC rates were notably lower at night (20.9% vs. 34.8%; *P = 0.01*), as was 30-day survival (47.0% vs. 59.3%; *P = 0.01*). EMS response times were significantly longer during nighttime hours (median and interquartile range: 12.4(7.4-14.6) vs. 11.2(6.2-13.5)(minutes); *P = 0.01*)

**Conclusions:** Patients with OHCA during off-hours were exposed to longer EMS response time as compared to procedures conducted during regular working hours. Furthermore, OHCA during night shift might be associated with a lower rate of ROSC and decreased 30-day survival

**What’s New?:** Out-of-hospital cardiac arrest (OHCA) remains a critical public health challenge with significant implications for morbidity and mortality. Despite advancements in emergency medical services systems and resuscitation techniques, survival rates following OHCA remain low, particularly during nighttime hours. This study provided a clinical view from a national perspective on the impact of day versus night shifts on the clinical outcomes in patients with OHCA. Propensity score match analysis was performed to evade risk of bias in the preselection process. This study suggested detrimental outcome in OHCA treatment during nighttime as compared to regular working hours. Patients from nighttime group were associated with longer response times as well as decreased rate of return of spontaneous circulation and 30-day survival as compared to daytime. These findings underscore the importance of systemic approach to improve OHCA outcome.

## Introduction

Out-of-hospital cardiac arrest (OHCA) remains a critical public health challenge with significant implications for morbidity and mortality. Despite advancements in emergency medical services (EMS) systems and resuscitation techniques, survival rates following OHCA remain low, particularly during nighttime hours ^1^. Previous research has highlighted differences in patient outcomes based on the time of day, with variations in return of spontaneous circulation (ROSC) and survival rates ^2–4^. Factors such as extended response times, reduced witnessed cardiopulmonary resuscitation (CPR) rates, and limited access to specialized care during nighttime hours have been shown to negatively impact outcomes ^4,5^. Moreover, workforce fatigue, resource limitations, and logistical challenges during night shifts may further contribute to the observed disparities^6^. Due to ethical constraints, randomized clinical trials in this setting are limited therefore, all-comers registries might serve as a valuable source of real-world data to address gaps in evidence. Thus, we sought to analyze the impact of day versus night shifts on OHCA outcomes in data from an unselected cohort of consecutive patients.

### Methods

This retrospective observational study collected data on cardiac arrest using mandatory electronic records maintained by emergency medical team personnel, including paramedics, nurses, and physicians. The study was conducted in Poland over a three-month period, with data gathered at the scene of each OHCA by emergency medical teams. Information regarding 30-day survival in follow up was gathered from the national death registry. The analysis focused on OHCA cases that occurred between September 1 and November 30, 2022. The patient flow chart is presented in Figure 1. All procedures were performed following local standards and Advance Life Support guidelines wherever applicable. All periprocedural complications were collected prospectively. All adverse events were diagnosed at the EMS team leader discretion in accordance with definitions in current ERC guidelines ^7^. However, data beyond the hospital discharge were not collected. The sample was specifically selected to enable comparisons with the multicenter EuReCa study ^8^. Patients were grouped by the time of their cardiac arrest using the Utstein framework ^9–11^. Data marked as “unknown” or “not recorded” were excluded. All ambulance service patients who experienced OHCA during the study period were included (n=7237). Propensity score matching (PSM) analysis was performed to match patients with cardiac arrest occurring during day to those with cardiac arrest occurred during night (On-hours: Monday to Sunday, 7:00 AM – 6:59 PM vs. Off-hours: Monday to Sunday, 7:00 PM – 6:59 AM). The stabilized weights were calculated using propensity scores obtained from a logistic regression model. The covariates included in the final propensity score model were: age, sex, CPR before EMS arrival, cause of cardiac arrest, location of cardiac arrest, presence of a witness on scene, shockable initial rhythm occurrence, and Automated External Defibrillator (AED) utilization. The nearest neighbor matching was performed. To minimize the standardized differences between groups, no match tolerance was applied. The 1:1 ratio was chosen to minimize bias without sacrificing test power in accordance with previous recommendations ^12^. Analyzed variables between groups included ROSC, 30-day survival, time of arrival, time on-scene and overall operation time. Continuous variables ware presented as median and interquartile range (IQR) for non-normally distributed variables, respectively. Categorical variables were presented as numbers and percentages. To check the normality of the distribution of the continuous variables we used the Shapiro-Wilk and the Kolmogorov-Smirnov with the Lilliefors correction tests. Comparison between groups was established using a Mann–Whitney U test for continuous variables and Chi-square test or Chi-square test with Fisher’s correction for categorical variables. For all inferential statistics statistical significance was defined as *P=0.05*. All calculations were done with SPSS 10.0 ® statistical software (SPSS Inc, Chicago IL, USA). Statistical criteria for selecting the optimal number of untreated subjects matched to each treated subject when using many-to-one matching on the propensity score. The study was approved by the institutional ethical board. The study was provided in accordance with ethical principles for clinical research based on the Declaration of Helsinki with later amendments.

**Figure 1.**
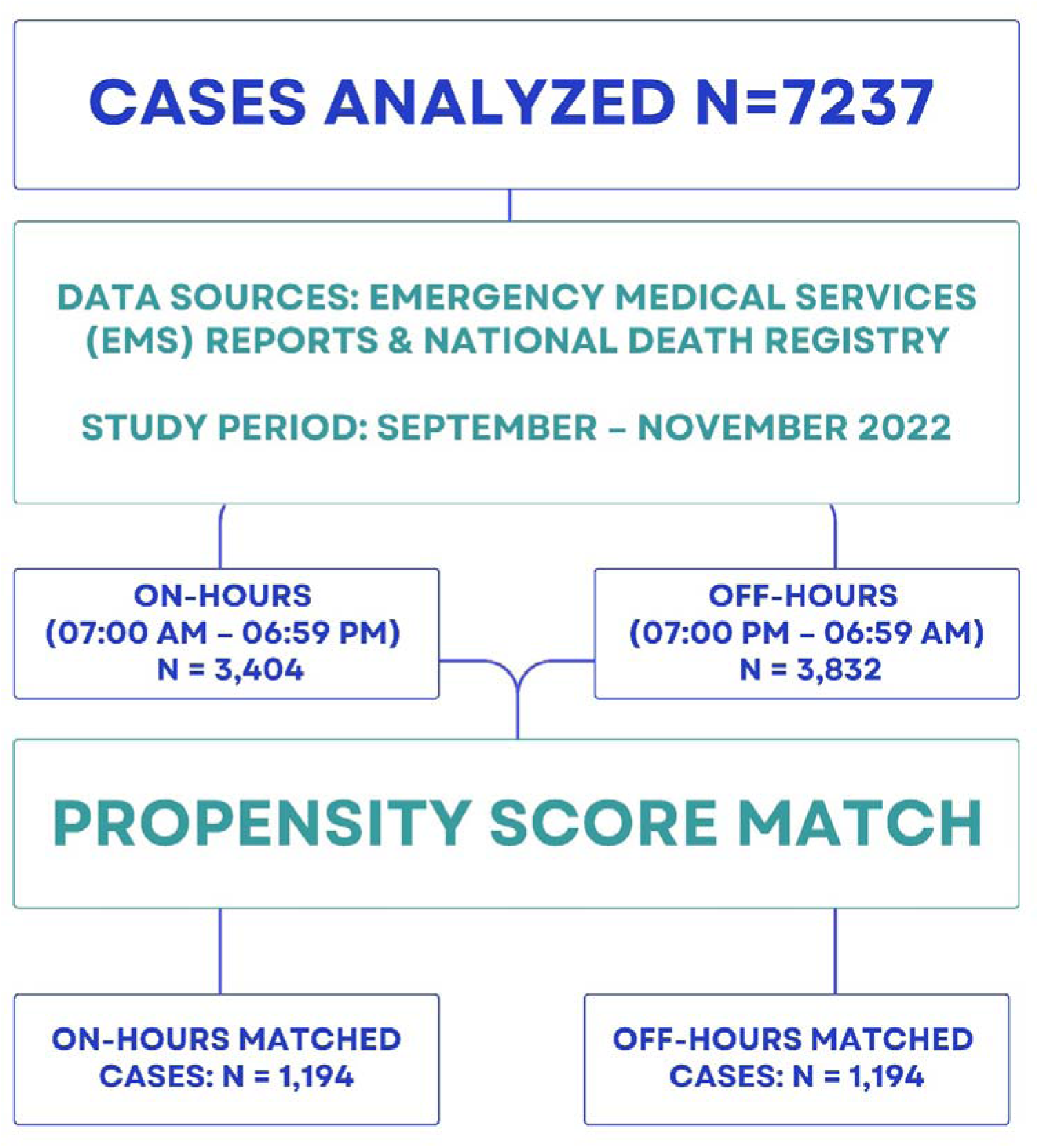
Study Population.

## Results

The characteristics of the study population are presented in Table 1. A total of 1194 matched pairs treated during day- and nighttime were evaluated. After PSM no significant differences in baseline patients’ characteristics were observed between groups (Table 2). The results obtained from this analysis are set out in Table 3, showing a significantly higher ROSC rate (34.8% vs. 20.9%; *P = 0.01*) and improved 30-day survival outcomes (59.3% vs. 47.0%; *P = 0.01*) during the day-as compared to nighttime. Patients in daytime group experienced shorter time to arrival (median and interquartile range (IQR):11.2(6.2-13.5) vs. 12.4(7.4-14.6) (minutes); *P = 0.01*) and longer on-scene times (median and IQR: 108.3(56.2-103.1) vs.107.1(59.9 – 106.7)(minutes); *P = 0.03*). Furthermore, overall operation time was found to be shorter at night 108.6 (65.0-113.5) vs (112.8(58.8-106.7) minutes; *P = 0.02*) longer on-scene times (108.2min vs. 107.1min; *P = 0.03*).

**Table 1.**
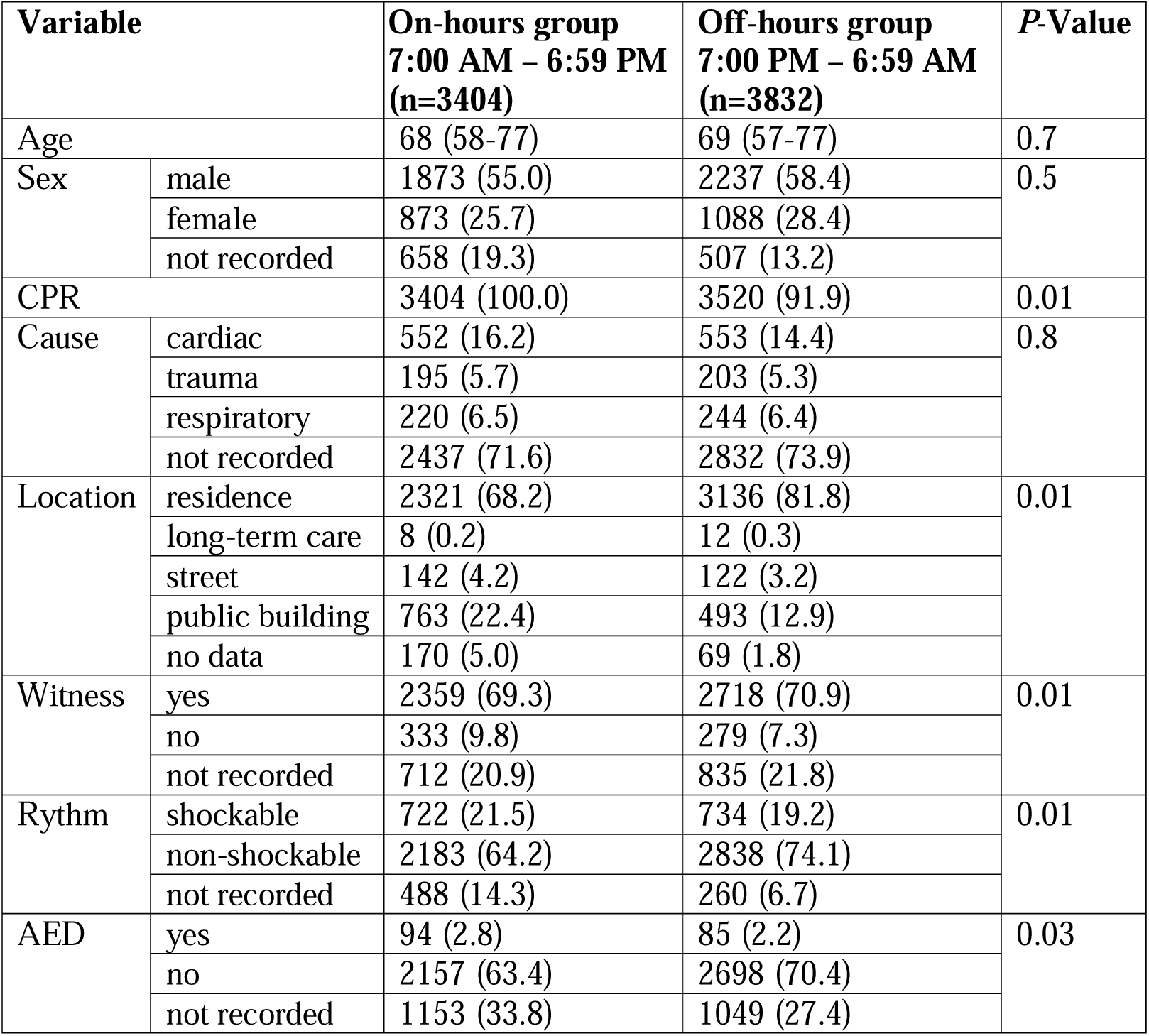
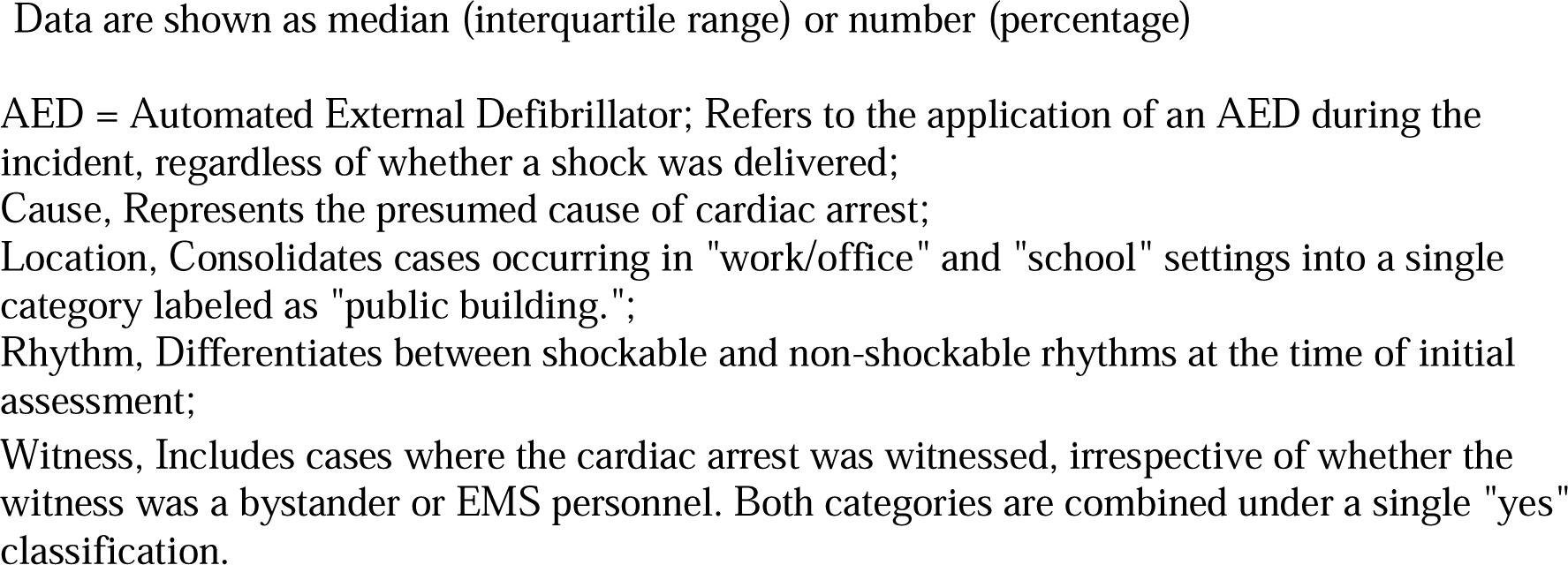
Characteristics of the study population before propensity score matching.

| Variable |  | On-hours group<br>7:00 AM – 6:59 PM<br>(n=3404) | Off-hours group<br>7:00 PM – 6:59 AM<br>(n=3832) | P-Value |
| --- | --- | --- | --- | --- |
| Age |  | 68 (58-77) | 69 (57-77) | 0.7 |
| Sex | male | 1873 (55.0) | 2237 (58.4) | 0.5 |
|  | female | 873 (25.7) | 1088 (28.4) |  |
|  | not recorded | 658 (19.3) | 507 (13.2) |  |
| CPR |  | 3404 (100.0) | 3520 (91.9) | 0.01 |
| Cause | cardiac | 552 (16.2) | 553 (14.4) | 0.8 |
|  | trauma | 195 (5.7) | 203 (5.3) |  |
|  | respiratory | 220 (6.5) | 244 (6.4) |  |
|  | not recorded | 2437 (71.6) | 2832 (73.9) |  |
| Location | residence | 2321 (68.2) | 3136 (81.8) | 0.01 |
|  | long-term care | 8 (0.2) | 12 (0.3) |  |
|  | street | 142 (4.2) | 122 (3.2) |  |
|  | public building | 763 (22.4) | 493 (12.9) |  |
|  | no data | 170 (5.0) | 69 (1.8) |  |
| Witness | yes | 2359 (69.3) | 2718 (70.9) | 0.01 |
|  | no | 333 (9.8) | 279 (7.3) |  |
|  | not recorded | 712 (20.9) | 835 (21.8) |  |
| Rythm | shockable | 722 (21.5) | 734 (19.2) | 0.01 |
|  | non-shockable | 2183 (64.2) | 2838 (74.1) |  |
|  | not recorded | 488 (14.3) | 260 (6.7) |  |
| AED | yes | 94 (2.8) | 85 (2.2) | 0.03 |
|  | no | 2157 (63.4) | 2698 (70.4) |  |
|  | not recorded | 1153 (33.8) | 1049 (27.4) |  |

Data are shown as median (interquartile range) or number (percentage)
AED = Automated External Defibrillator; Refers to the application of an AED during the incident, regardless of whether a shock was delivered;

**Table 2.**
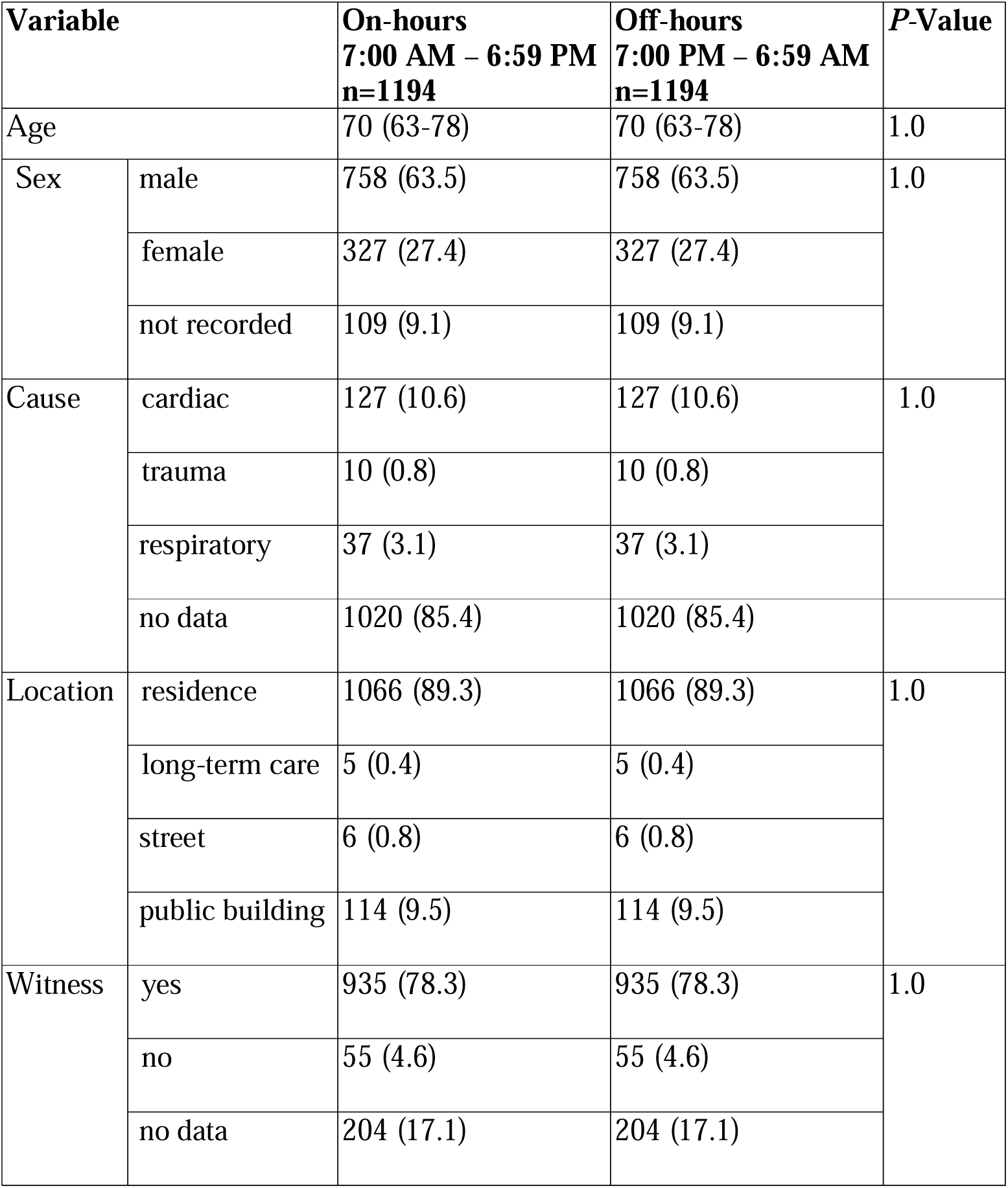
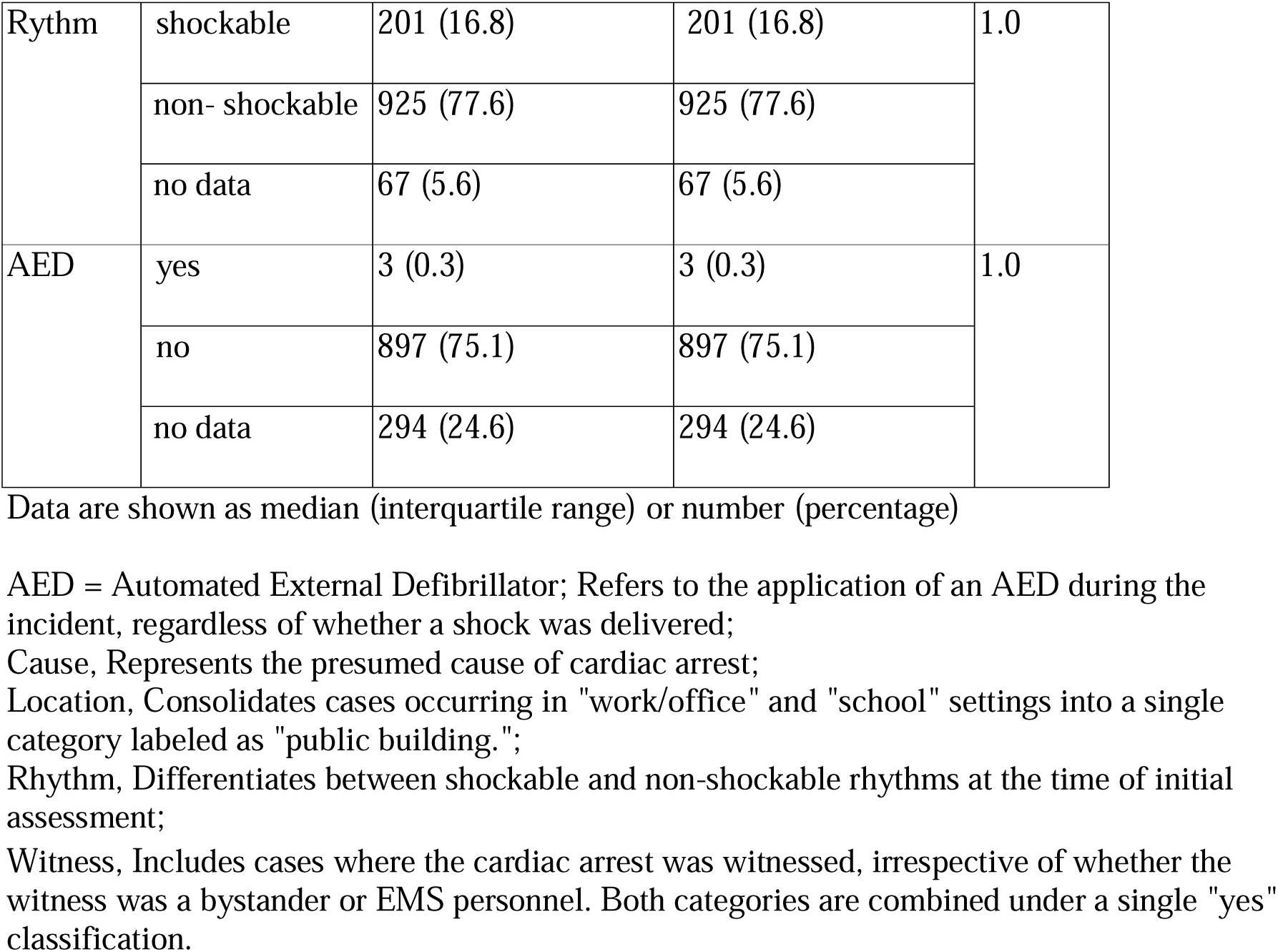
Baseline characteristics of the study population after propensity score matching.

**Table 3.**
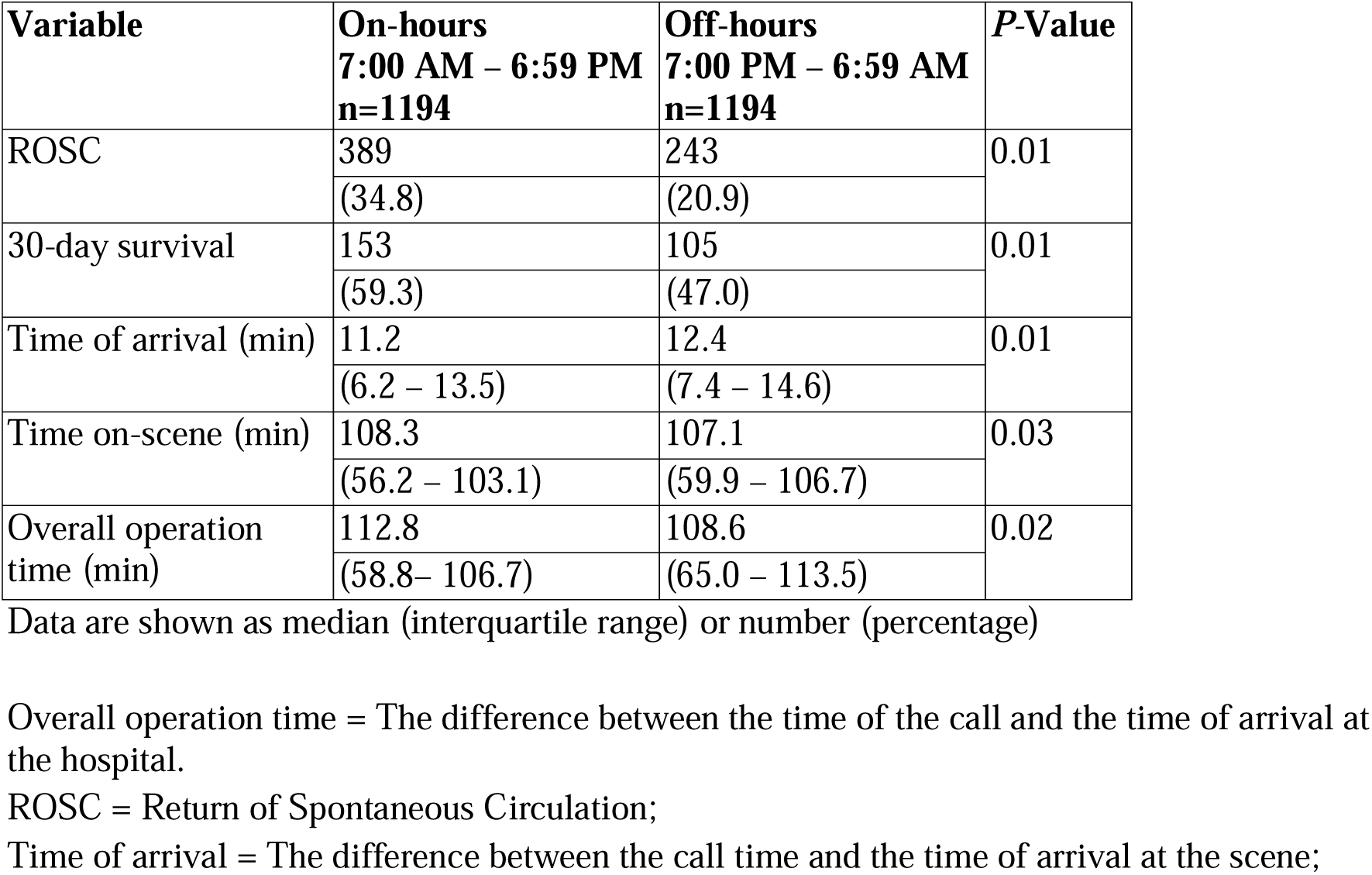
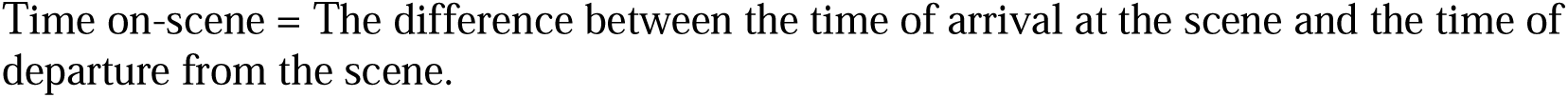
Comparison of OHCA outcomes and response times between groups.

## Limitations

While our findings provide valuable insights, there are certain limitations that must be acknowledged. The most important is the nonrandomized design with all related bias. The risk of confounding factors cannot be excluded. However, a PSM calculation was provided to imitate randomization procedure and evade risk of bias in the preselection process. The presented analysis has not included all patients with OHCA during the study period. Some patients with highest risk burden and the most severe condition might die without EMS call, thus they were not included in the database. There are notable gaps in data completeness: while ROSC data is available for nearly all patients, 30-day survival data is limited to about 250 pairs of matched patients. This discrepancy could affect the robustness of our findings. The analysis relies on paramedic reports for ROSC and the national death registry for 30-day survival rates, initially connected by PESEL number and anonymized before analysis. Any ROSC is marked in paramedic reports, while 30-day survival was chosen for its standardized timeline from the Utstein framework^10^. The use of the PESEL number to connect paramedic reports and National Death Registry to monitor deaths excluded some patients who had not yet been assigned identification numbers, resulting in missing data and underrepresentation of cases in terms of 30-day survival. PESEL is a unique 11-digit national identification number assigned to all Polish citizens and residents, used for identity verification in administrative, legal, and medical systems. This limitation impacts the completeness and accuracy of survival data, potentially affecting the study’s conclusions. There might be additional risk of patients overlapping across the cohorts, mainly in those registered between shifts. Furthermore, senior staff availability and experience of medical team might be considered as possible alterable aspects influencing outcomes. Despite all these described limitations, our study presented experience from a large, unselected cohort of patients; thus, outcomes might be adapted to the general population.

## Discussion

Presented results suggested detrimental outcome in OHCA treatment during nighttime as compared to regular working hours. Nighttime group was associated with longer response times as well as decreased rate of ROSC and 30-day survival as compared to daytime. These results are in line with previous research and underscore the critical importance of timely responses in OHCA management ^13–17^. Multiple factors likely contribute to this disparity, including workforce fatigue, variations in available resources like dispatcher assistance ^18,19^, medical staff competence and the extent of witness involvement ^15,20^. The findings of Bartlett at all. (2022) described how sleepiness effected at EMS work at night ^21^. Reaction time is longer, cognitive decisions are harder to make, and more mistakes occur. Furthermore, the presence of specialized hospital care in emergency department and personnel during the day likely increases survival outcomes, whereas nighttime care may encounter challenges such as limited staffing, specialist availability and diagnostic support ^22^. Variations in response time might also influence the outcome. In terms of on-scene time, the slight reduction at night (108.2min vs.107.1min; *P = 0.03*) suggested different treatment strategies. Daytime operations might allow for more comprehensive on-site interventions due to better resource availability and presence of more experienced medical staff. While reduced traffic congestion at night might appear advantageous, the reality of navigation difficulties, and possible delays in dispatch can counteract these benefits, resulting in longer response intervals. This study demonstrated that EMS operations during night shifts are less effective, emphasizing the need for systemic changes to improve after-hours response capabilities. According to the Central Statistical Office (Główny Urząd Statystyczny), as of December 31, 2023, emergency medical teams in Poland consisted of nearly 12,900 personnel. Paramedics represented the largest group, with over 11,200 members, followed by more than 1,000 emergency medical system nurses, over 300 physicians, and nearly 300 other staff members ^23^. The operational structure of EMS in Poland characterized working under a 7-7 shift system, alternating day and night shifts. The standard daily working time for EMS personnel on a full-time contract is 7 hours and 35 minutes. However, the equivalent working time system is commonly used in practice, offering greater flexibility in scheduling while ensuring compliance with the statutory weekly working limits. Following each night shift, a minimum rest period of 11 hours is required, whereas after a 24-hour shift, a minimum of 24 hours of rest must be provided. However, these regulations do not apply to B2B contracts, which are commonly chosen by a significant number of EMS personnel. A more stable work environment, regulated shifts, and standardized rest periods could improve overall system effectiveness and response quality. In the United Kingdom, EMS personnel work under a variety of shift patterns to ensure continuous 24/7 service, tailored to operational demands, environmental factors, and resource availability. One of the most commonly used schedules is the “2-2-4” system, where staff complete two consecutive 12-hour day shifts (e.g., 06:00–18:00), in accordance with the guidelines outlined by the Health and Safety Executive in *Managing Shiftwork – Health and Safety Guidance* ^24^. The B2B contract offers flexibility in choosing shift rotations, allowing individuals to tailor their work schedules based on personal preferences ^25^. However, it is worth considering established guidelines, such as those provided by the American College of Emergency Physicians. Their key recommendations suggest implementing a forward-rotating shift schedule (day to evening to night), managing shift lengths and consecutive work periods to prevent fatigue, and structuring night shifts either as isolated shifts or in longer blocks spanning several weeks ^25^. Furthermore, witnessed CPR might improve survival in cardiac arrest ^15,20^. However, available data revealed that bystander CPR is infrequent, particularly at night, further exacerbating poor outcomes. Implementing a national first responder system, expanding public CPR training programs, and increasing access to AEDs could allow to close the gap in outcomes between day and night ^26,27^. Despite longer overall operation time during daytime both mortality and ROSC rates seem to be beneficial as compared to outcomes from night shift. Since there was no difference in baseline characteristics presented results cannot be easily explained by impact of bystander or prevalence of shockable rhythm and AED utilization. This outcome might by partialy elucidated by lower quality of chest compression/ALS manoeuvers and general fatigue of medical staff at night. In addition, level of experience and dexterity in ALS procedures might have impact on this outcome. However, such data was not collected in this registry. Furthermore, hesitation in ambulance call especially in older population might result in more aggravated condition and higher risk burden of cardiac arrest. Previously published studies suggested diurnal variability in myocardial perfusion, particularly early in the morning and peak in platelet aggregation during night hours ^28^. Both patient-related and systemic factors might be responsible for presented phenomenon. These findings underscore the importance of systemic approach ^29^ to improve OHCA outcome. Addressing workforce issues such as implementing more sustainable work schedules, strengthening dispatcher-assisted CPR efforts, Public-Access Defibrillation and public education on AED usage, particularly during nighttime hours, could further support early intervention. Additionally, ensuring hospital readiness and coordination at night would help minimize delays and improve patient outcomes. Patients experiencing OHCA during off-hours faced longer EMS response time as compared to procedures conducted during regular working hours. Furthermore, OHCA occurring during night shift might be associated with a lower rate of ROSC and decreased 30-day survival as compared to patients treated during daytime.

## Author Contributions (CRediT)

- *Anna* Żą*dło:* Conceptualization, Methodology, Investigation, Data Curation, Writing – Original Draft
- *Grzegorz Cebula:* Conceptualization, Investigation, Data Curation
- *Monika Bednarek-Chałuda:* Writing – Review & Editing, Data Curation
- *Izabela A. Karpi*ń*ska:* Methodology, Investigation, Data Curation, Writing – Original Draft
- *Tomasz Tokarek:* Conceptualization, Methodology, Investigation, Data Curation, Writing – Review & Editing

## Conflict of Interes1t

None declared.

## Ethics Approval Statement

Approved by the Bioethics Committee for Research Studies of the Jagiellonian University Medical College (approval no. 118.0043.1.299.2024) on September 26, 2024.

## Data Availability

All data produced in the present study are available upon reasonable request to the authors

## References

1. Berdowski J, Berg RA, Tijssen JGP, Koster RW. Global incidences of out-of-hospital cardiac arrest and survival rates: Systematic review of 67 prospective studies. Resuscitation. 2010; 81: 1479–1487. Doi:10.1016/j.resuscitation.2010.08.006

2. Koike S, Tanabe S, Ogawa T, et al. Effect of time and day of admission on 1-month survival and neurologically favourable 1-month survival in out-of-hospital cardiopulmonary arrest patients. Resuscitation. 2011; 82: 863–868. Doi:10.1016/j.resuscitation.2011.02.007

3. Lin P, Shi F, Wang L, Liang ZA. Nighttime is associated with decreased survival for out of hospital cardiac arrests: A meta-analysis of observational studies. Am J Emerg Med. 2019;37:524–529. Doi:10.1016/j.ajem.2019.01.004

4. Bagai A, McNally BF, Al-Khatib SM, et al. Temporal Differences in Out-of-Hospital Cardiac Arrest Incidence and Survival. Circulation (Baltimore). 2013;128:2595–2602. Doi:10.1161/circulationaha.113.004164

5. Daya MR, Schmicker RH, Zive DM, et al. Out-of-hospital cardiac arrest survival improving over time: Results from the Resuscitation Outcomes Consortium (ROC). Resuscitation. 2015;91:108–115. Doi:10.1016/j.resuscitation.2015.02.003

6. Marvin G, Schram B, Orr R, Canetti EFD. Occupation-Induced Fatigue and Impacts on Emergency First Responders: A Systematic Review. Published online 2023;7055 Doi:10.3390/ijerph20227055

7. Perkins GD, Graesner JT, Semeraro F, et al. European Resuscitation Council Guidelines 2021: Executive summary. Resuscitation. 2021;161:1–60. Doi:10.1016/j.resuscitation.2021.02.003

8. Maurer H, Masterson S, Tjelmeland IBM, et al. Eureca – The European Registry of Cardiac Arrest and the related studies. Resusc Plus. 2024;19. Doi:10.1016/j.resplu.2024.100666

9. Idris AH, Bierens JJLM, Perkins GD, et al. 2015 revised Utstein-style recommended guidelines for uniform reporting of data from drowning-related resuscitation: An ILCOR advisory statement. Resuscitation. 2017;118:147–158. Doi:10.1016/j.resuscitation.2017.05.028

10. Grasner JT, Bray JE, Nolan JP, et al. Cardiac arrest and cardiopulmonary resuscitation outcome reports: 2024 update of the Utstein Out-of-Hospital Cardiac Arrest Registry template. Resuscitation. 2024; 27:203–223. Doi:10.1016/j.resuscitation.2024.110288

11. Cummins RO, Chamberlain DA, Abramson NS, et al. Recommended guidelines for uniform reporting of data from out-of-hospital cardiac arrest: the Utstein Style. A statement for health professionals from a task force of the American Heart Association, the European Resuscitation Council, the Heart and Stroke Foundation of Canada, and the Australian Resuscitation Council. Circulation. 1991;84:960–975. Doi:10.1161/01.cir.84.2.960

12. Njei B, McCarty TR, Sharma P, et al. Bariatric Surgery and Hepatocellular Carcinoma: A Propensity Score Matched Analysis. Obes Surg. 2018;28:3880. Doi:10.1007/S11695-018-3431-5

13. Nichol G, Thomas E, Callaway CW, et al. Regional Variation in Out-of-Hospital Cardiac Arrest Incidence and Outcome. [published correction appears in JAMA. 2008 Oct 15;300:1763]. JAMA. 2008;300:1423–1431. Doi:10.1001/jama.300.12.1423

14. Ong MEH, Shin S Do, De Souza NNA, et al. Outcomes for out-of-hospital cardiac arrests across 7 countries in Asia: The Pan Asian Resuscitation Outcomes Study (PAROS). Resuscitation. 2015;96:100–108. Doi:10.1016/j.resuscitation.2015.07.026

15. Važanić D, Prkačin I, Nesek-Adam V, et al. Out-of-hospital cardiac arrest outcomes-bystander cardiopulmonary resuscitation rate improvement. Acta clin croat. 2022; 61: 265–272.

16. Rudner R, Jalowiecki P, Karpel E, et.al. Survival after out-of-hospital cardiac arrests in Katowice (Poland): Outcome report according to the “Utstein style.” Resuscitation. 2004;61:315–325. Doi:10.1016/j.resuscitation.2004.01.020

17. Gräsner JT, Lefering R, Koster RW, et al. Eureca ONE—27 Nations, ONE Europe, ONE Registry: A prospective one month analysis of out-of-hospital cardiac arrest outcomes in 27 countries in Europe. Resuscitation. 2016;105:188–195. Doi:10.1016/j.resuscitation.2016.06.004

18. Bagheri S, Sadeghi SM, Kazemi T, Nadimi E. Dispatcher-Assisted Bystander Cardiopulmonary Resuscitation (Telephone-CPR) and Outcomes after Out of Hospital Cardiac Arrest. Trauma Research Center, Shiraz University of Medical Sciences Seyed Bagheri SM et al Bull Emerg Trauma. 2019;7:307–313. Doi:10.29252/beat-0703015

19. Xu J, Qu M, Dong X, et al. Tele-Instruction Tool for Multiple Lay Responders Providing Cardiopulmonary Resuscitation in Telehealth Emergency Dispatch Services: Mixed Methods Study. J Med Internet Res. 2023;25. Doi:10.2196/46092

20. Liou FY, Lin KC, Chien CS, et al. The impact of bystander cardiopulmonary resuscitation on patients with out-of-hospital cardiac arrests. Journal of the Chinese Medical Association. 2021;84:1078–1083. Doi:10.1097/jcma.0000000000000630

21. Bartlett D, Hansen S, Cruickshank T, et al. Effects of sleepiness on clinical decision making among paramedic students: A simulated night shift study. Emergency Medicine Journal. 2022;39:45–51. Doi:10.1136/emermed-2019-209211

22. Drennan J, Murphy A, mccarthy VJC, et al. The association between nurse staffing and quality of care in emergency departments: A systematic review. Int J Nurs Stud. 2024;153. Doi:10.1016/j.ijnurstu.2024.104706

23. Statistics Poland. Emergency medical aid and rescue services in 2023. Warsaw: Statistics Poland; 2024 [cited 2025 Mar 28]. Available from: https://stat.gov.pl/obszary-tematyczne/zdrowie/zdrowie/pomoc-dorazna-i-ratownictwo-medyczne-w-2023-roku,14,8.html

24. Managing Shiftwork_J: Health and Safety Guidance. Health and Safety Executive; 2009.

25. Rischall ML, Chung AS, Tabatabai R, Doty C, Hart D. Emergency Medicine Resident Shift Work Preferences: A Comparison of Resident Scheduling Preferences and Recommended Schedule Design for Shift Workers. AEM Educ Train. 2018;2:229. Doi:10.1002/aet2.10104

26. Telec W, Baszko A, Dąbrowski M, et al. Automated external defibrillator use in public places: A study of acquisition time. Kardiol Pol. 2018;76:181–185. Doi:10.5603/kp.a2017.0199

27. Żuratyński P, Ślęzak D, Dąbrowski S, et.al. Use of public automated external defibrillators in out-of-hospital cardiac arrest in Poland. Medicina (Lithuania). 2021;57:298. Doi:10.3390/medicina57030298

28. Tokarek T, Dziewierz A, Plens K, et al. Percutaneous coronary intervention during on- and off-hours in patients with ST-segment elevation myocardial infarction. Hellenic Journal of Cardiology. 2021;62:212–218. Doi:10.1016/j.hjc.2021.01.011

29. Marijon E, Narayanan K, Smith K, et al. The Lancet Commission to reduce the global burden of sudden cardiac death: a call for multidisciplinary action. The Lancet. 2023;402:883–936. Doi:10.1016/S0140-6736(23)00875-9

